# Long-term Exposure to PM_2.5_ Components and Lipid Profiles in World Trade Center Health Program General Responders

**DOI:** 10.64898/2026.06.10.26355272

**Authors:** Helena Krasnov, Pablo Knobel, Leon Hsiao-Hsien Hsu, Susan L. Teitelbaum, Mary Ann McLaughlin, Allan C. Just, Itai Kloog, Maayan Yitshak Sade

## Abstract

Previous studies have shown that fine particulate matter (PM_2.5_) is associated with elevated blood lipids, but very few studies have examined the associations with specific constituents of PM_2.5_. We studied the associations between exposure to annual PM_2.5_ and its 14 constituents, and repeated blood lipid measurements among general responders enrolled in the World Trade Center Health Program between 2003 and 2019 (*n* = 44,876). We used generalized additive mixed effect models to investigate the single-pollutant associations with repeated measures of blood total cholesterol (TC), and low-density lipoprotein (LDL-C) levels. We then used linear generalized weighted quantile sum regression with a random intercept for participant ID to account for the clustering of repeated measures and evaluate the combined associations with the component mixture. A decile increase in the mixture of 14 PM_2.5_ chemical components was associated with 0.375 mg/dL increase in TC levels (95% confidence Interval (CI): 0.174-0.577) and 0.302 mg/dL increase in LDL-C (95% CI: 0.063, 0.540). Lead, organic carbon, and iron were major drivers of both associations. Component-specific models also show higher TC and LDL-C levels associated with interquartile range increases in organic carbon (0.472, [95% CI: 0.027, 0.918] and 0.648 [95% CI: 0.136, 1.160]) and iron exposure (1.081, [95% CI: 0.630, 1.532] and 0.748 [95% CI 0.318, 1.178]). Effects are more pronounced among responders with higher World Trade Center exposures, suggesting increased vulnerability.

These findings are highly significant for a population exposed to extreme air pollution event and susceptible to lipid alterations that might trigger cardiovascular events.

## 1. Introduction

General responders involved in post-9/11 emergency response activities at the World Trade Center (WTC) were exposed to severe and complex chemical exposures not experienced by the general population. Previous studies demonstrated that exposure to these toxicants is associated with numerous long-term cardiovascular outcomes among responders ^1,2^ and may have increased responders’ susceptibility to subsequent air pollution exposures, particularly fine particulate matter (PM_2.5_), later in life ^3^. Although toxicological studies have proposed pathways such as oxidative stress and systemic inflammation, the mechanisms linking PM_2.5_ and cardiovascular outcomes are still not fully elucidated ^4^. Emerging evidence indicates that lipid profile, namely Total Cholesterol (TC), and Low-Density Lipoprotein Cholesterol (LDL-C) levels may play a role on the pathway linking exposure to PM_2.5_ with cardiovascular disease (CVD) risk ^5^. Further large-scale epidemiological studies are needed to support the link between exposure to PM_2.5_ and lipid levels.

The association between PM_2.5_ and cholesterol was recently reported in general responders enrolled in the World Trade Center Health Program (WTCHP) ^2^, a federal program that provides medical monitoring and treatment for certified physical and mental health conditions for eligible responders. While this study adds robust evidence, it does not address the complexity of PM_2.5_ as a mixture with substantial heterogeneity in physical characteristics, chemical composition, and sources. To date, few studies investigated the associations between PM_2.5_ constituents and lipid profiles, with the majority relying on single-pollutant models ^6,7^. This approach fails to account for multicollinearity with co-pollutants, potentially leading to confounding and biased effect estimates. Exposure mixture approaches, overcome this limitation by accounting for the health effects of concurrent, and often correlated, exposures.

Building on a prior study which showed differential associations of source-apportioned PM_2.5_ exposures and glycemic status among WTCHP general responders ^3^, this study examines the associations of specific constituents of PM_2.5_ with repeated measurements of TC, and LDL-C levels among the WTCHP general respondents cohort (GRC). Additionally, we study whether these associations are modified by WTC-related exposures.

## 2. Methods

### 2.1 Study population and outcomes

We included all available laboratory samples collected from WTCHP general responders between the years 2003 to 2019 who enrolled in the WTC Health Program and were offered annual monitoring visits ^8^, during which a healthcare professional collected laboratory samples along with a survey of demographic and health-related questions. Laboratory results were performed by Mount Sinai, NYU, Stony Brook, and the Northwell Clinical Centers of Excellence. Sociodemographic information was collected at the first visit, and address histories were updated at each monitoring visit starting 2012. We included repeated measurements of TC (n = 143,555), and LDL-C (n = 96,759) obtained during the monitoring visits from 44,876 members of the WTCHP GRC.

### 2.2 Exposure

We obtained annual PM_2.5_ mass estimates from our hybrid prediction models using extreme gradient boosting (XGBoost) and an Inverse Distance Weighting interpolation (IDW) Synthesis (the XIS model) ^9^, combined with satellite-derived aerosol optical depth among twenty other topographical, land use and meteorological predictors, resulting in our ability to reconstruct address-specific daily PM_2.5_ estimates for each responder across the contiguous United States (CONUS) during the period from 2003 through 2023. The model is trained and cross-validated using PM_2.5_ measurements from the U.S. Environmental Protection Agency Air Quality System, with excellent model performance. The nationally mean absolute error of daily predictions is 2.09 _μ_g/m3, which is a lower error in more recent years and in the South and Northeast regions. We additionally obtained annual PM_2.5_ chemical component estimates from our component models, generating mean concentrations of 14 PM components on a 50m resolution grid in urban areas across the U.S ^10^. The analyzed components included bromine - Br; calcium - Ca; copper - Cu; iron - Fe; potassium - K; nickel - Ni; lead - Pb; silicon - Si; vanadium - V; zinc - Zn; elemental carbon – EC; nitrate - NO_3_; organic carbon - OC; and sulfate - SO_4_. The models incorporate measurements from air pollution and weather monitoring sites, and over 160 predictor variables including land use information, meteorological covariates, and satellite observations. Predictions from three machine learning algorithms were integrated using super-learning and ensemble weighted-averaging models. The R^2^ values in an unseen test set were above 0.90 on average.

The annual exposures were linked to geocoded residential addresses. Using the annual addresses and reported moving dates, we constructed a longitudinal dataset with annual observations, each linked to the most recently reported geocoded address; for years prior to 2012, we assigned the closest reported address. Among movers, if a responder moved mid-year, the average value of both locations was calculated.

### 2.3 Covariates

We adjusted the models for individual characteristics, including age, sex (male and female) and race (Black, White, other and unknown). Additionally, we adjusted the models for census tract-level socioeconomic variables used as a proxy for sociodemographic neighborhood adjustment: percentage of the population below poverty line (annual income < $11,484), percentage of Black population, percentage with no high school diploma, and median household income. Finally, we adjusted the models for annual average temperature exposure, using temperature estimates from a high-resolution geospatial model ^11^, monitoring visit year, and residential region (Midwest, Northeast, Southeast, Southwest, West) to account for regional differences around the US.

### 2.4 Statistical analysis

First, we used generalized additive linear mixed effect models to investigate the associations of PM_2.5_ mass and the individual PM_2.5_ constituents exposures with repeated measures of blood TC, and LDL-C levels. Results are reported as percentage change in lipid levels.

We winsorized the exposures to the 0.5% and 99.5% of the exposure distributions to avoid bias due to extreme values. We tested for potential non-linear associations using penalized spline functions for the exposures. Since all the associations were linear or nearly linear (Figure S1), we used linear exposure terms across all the models. All models included a random intercept for each responder to account for repeated measures. In addition to the covariates described above, models were adjusted for a penalized spline for the year of testing to control for the time trend. The single-pollutant models were additionally adjusted for total PM_2.5_ mass. For the components that were highly correlated (r > 0.80) with PM_2.5_ (V, SO_4_, Figure S2) we employed a residualization approach to isolate the effect of the component independent of total PM_2.5_ mass and avoid confounding by the overall air pollution burden ^12^. Total PM_2.5_ mass was first regressed on that component. The residuals representing the portion of PM_2.5_ mass not explained by the component were then included as a covariate in the primary model.

Next, we used linear generalized weighted quantile sum (gWQS) regression to evaluate the combined effect of the component mixture and their relative weight on the lab results ^13^. The dataset is randomly split into training (40%) and validation (60%) sets. In the training set, the components are standardized and ranked into increments and are combined into a weighted additive index, with weights constrained to be positive or negative with a sum of one. Larger weights indicate greater contribution to the mixture effect, while near-zero weights indicate minimal impact. To address high intercorrelation and improve weight stability, 100 bootstrap samples were drawn from the training set. We selected deciles (q=10) based on the lowest Bayesian Information Criterion (BIC) values after comparing the results of 4 to 10 increments. We incorporated a random intercept for participant ID to account for the clustering of repeated measures and we adjusted the models for individual and census tract-level characteristics, annual average temperature, monitoring visit year, and residential region. To evaluate the robustness of our findings, we repeated the models using 8 and 9 quantiles instead of the primary specification. Finally, WTC exposure-stratified analysis was conducted to investigate potentially differential susceptibility between exposure groups, including differences according to exposure level, arrival time at the site, and loss of someone. Statistical significance for effect modification was determined from the interaction term in the full gWQS regression model.

## 3. Results

We included a total of 482,618 person years in the analysis. This longitudinal dataset comprised 143,555 TC measurements and 96,759 LDL-C measurements from 44,876 responders (Table 1). The mean age of the responders was 50.88, while the average age of participants providing TC samples was approximately 52 years. The mean age of the responders LDL-C lab samples throughout the study period was approximately 54 years. The majority of the responders were white (60%), and 85% were male. Mean sample value for TC was 194.94 mg/dL and 111.21 mg/dL for LDL-C. Most of the responders (77.81%) reside in the northeast of the US. The summary statistics of the PM_2.5_ chemical components levels evaluated in this study are summarized in Table 2. The correlations between the component exposures are presented in Figure S2. The most substantial correlations were observed among PM_2.5_ and SO_4_ (r = 0.89), PM_2.5_ and V (r = 0.85), Zn and EC (r = 0.85), Fe and EC (r = 0.82), and V and SO_4_ (r = 0.84). Interquartile range (IQR) increases in annual PM_2.5_ exposure were associated with a 1.294 mg/dL increase in TC levels (95% CI: 0.579, 2.010) and 0.827 mg/dL increase in LDL-C (95% CI: 0.352, 1.303). In the single-component models, adjusted for PM_2.5_ mass, we found increases in TC and LDL-C levels associated with Ca, Cu, V, and OC exposure. Higher TC levels were also associated with Zn, EC, and NO_3_ exposures while high LDL-C was associated with Ni exposure (Table 3).

**Table 1.** Clinical lab results population characteristics,N = 44,876 (2003-2019)

| Variable | Overall<br>(N= 482,618) | TC<br>(N = 143,555) | LDL-C<br>(N = 96,759) |
| --- | --- | --- | --- |
| Sociodemographic characteristics, N (%) |  |  |  |
| Age, Mean (SD) | 50.88 (9.75) | 51.74 (9.56) | 54.15 (8.71) |
| Race |  |  |  |
| *Black* | 51,093 (10.59%) | 16,818 (11.72%) | 10,921 (11.29%) |
| *White* | 290,005 (60.09%) | 80,290 (55.93%) | 54,213 (56.03%) |
| *Other* | 64,527 (13.37%) | 26,237 (18.28%) | 17,338 (17.92%) |
| *Unknown* | 76,993 (15.95%) | 12,210 (14.08%) | 14,287 (14.77%) |
| Highest Education Level |  |  |  |
| *Elementary School* | 36,409 (7.54%) | 11,434 (7.96%) | 6,979 (7.21%) |
| *High School* | 106,147 (21.99%) | 29,804 (20.76%) | 18,752 (19.38%) |
| *College* | 130,669 (27.08%) | 38,228 (26.63%) | 26,052 (26.92%) |
| *Other*<sup>a</sup> | 189,605 (39.29%) | 58,866 (41.01%) | 41,276 (42.66%) |
| *Unknown* | 19,788 (4.10%) | 5,223 (3.6%) | 3,700 (3.8%) |
| Sex |  |  |  |
| *Male* | 414,022 (85.79%) | 121,973 (84.97%) | 82,398 (85.16%) |
| *Female* | 68,596 (14.21%) | 21,581 (15.03%) | 14,361 (14.84%) |
| Neighborhood characteristics,<br>Mean (SD) |  |  |  |
| % Black population | 0.93% (1.98) | 1.15% (2.17) | 1.14% (2.16) |
| % Below the poverty line | 9.69% (9.04) | 10.71% (9.73) | 10.06% (9.16) |
| % No high school education | 3.99% (4.02) | 4.51% (4.40) | 4.06% (3.96) |
| Median household income | 82,445 (33,785) | 83,221 (35,147) | 89,748 (36,082) |
| Outcome, Mean (SD) | (---) | 194.94 (38.86) | 111.21 (33.64) |
| Region |  |  |  |
| Midwest | 761 (0.16%) | 109 (0.08 %) | 44 (0.05 %) |
| Northeast | 375,509 (77.81%) | 130,127 (90.65 %) | 85,967 (88.85 %) |
| South-East | 19,780 (4.10%) | 3,278 (2.28 %) | 2,192 (2.27 %) |
| South-West | 934 (0.19%) | 125 (0.09 %) | 73 (0.08 %) |
| West | 2,748 (0.57%) | 374 (0.26 %) | 212 (0.22 %) |
| Unknown | 82, 886 (17.2%) | 9,542 (6.6%) | 8,271 (8.5%) |

*Demographic and clinical characteristics are summarized at the laboratory sample level. Data are presented as n (%) unless otherwise indicated. Percentages are based on available data for each characteristic a Professional School/BA/BS/Associate DegreeSD = Standard Deviation; TC= Total Cholesterol; LDL-C = low-density lipoprotein*

**Table 2.** Summary statistics of main air pollutant exposures (2003-2019)

| Exposure | Mean (SD) | IQR |
| --- | --- | --- |
| Total PM<sub>2.5</sub> (µg/m<sup>3</sup>) | 8.39 (2.14) | 2.90 |
| PM<sub>2.5</sub> components |  |  |
| Br (ng/m<sup>3</sup>) | 2.57 (0.85) | 0.96 |
| Ca (ng/m<sup>3</sup>) | 35.24 (12.75) | 14.20 |
| Cu (ng/m<sup>3</sup>) | 3.87 (1.56) | 2.045 |
| Fe (ng/m<sup>3</sup>) | 71.84 (23.44) | 28.93 |
| K (ng/m<sup>3</sup>) | 49.08 (7.82) | 7.61 |
| Ni (ng/m<sup>3</sup>) | 1.53 (1.17) | 0.82 |
| Pb (ng/m<sup>3</sup>) | 2.94 (1.60) | 3.17 |
| Si (ng/m<sup>3</sup>) | 62.68 (25.29) | 17.46 |
| V (ng/m<sup>3</sup>) | 1.19 (1.15) | 1.34 |
| Zn (ng/m<sup>3</sup>) | 11.39 (4.80) | 5.72 |
| EC (µg/m<sup>3</sup>) | 0.74 (0.29) | 0.33 |
| NO<sub>3</sub> (µg/m<sup>3</sup>) | 1.12 (0.38) | 0.47 |
| OC ($\mu\text{g}/\text{m}^3$) | 1.77 (0.33) | 0.42 |
| SO<sub>4</sub> ($\mu\text{g}/\text{m}^3$) | 1.85 (0.93) | 1.33 |

*IQR = Interquartile Range; SD = Standard Deviation; PM<sub>2.5</sub> = Fine Particulate Matter.*
*Br - Bromine; Ca - Calcium carbonate; Cu - Copper; Fe - Iron; K - Potassium; Ni - Nickel; Pb - Lead; Si - Silicon; V - Vanadium; Zn - Zinc; EC - Elemental Carbon; NO<sub>3</sub> - Nitrate; OC - Organic Carbon; SO<sub>4</sub> - Sulfate.*

**Table 3.**
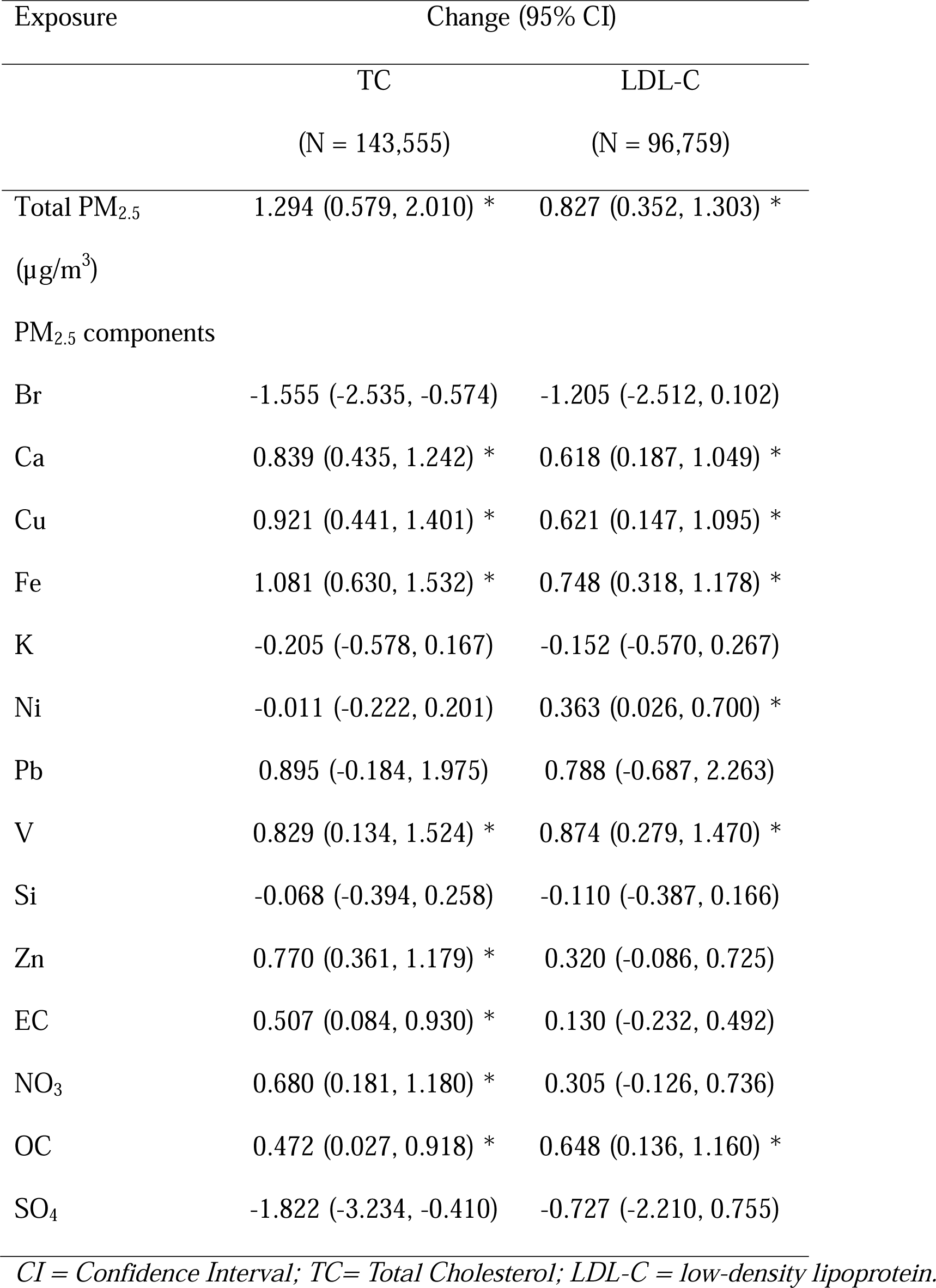
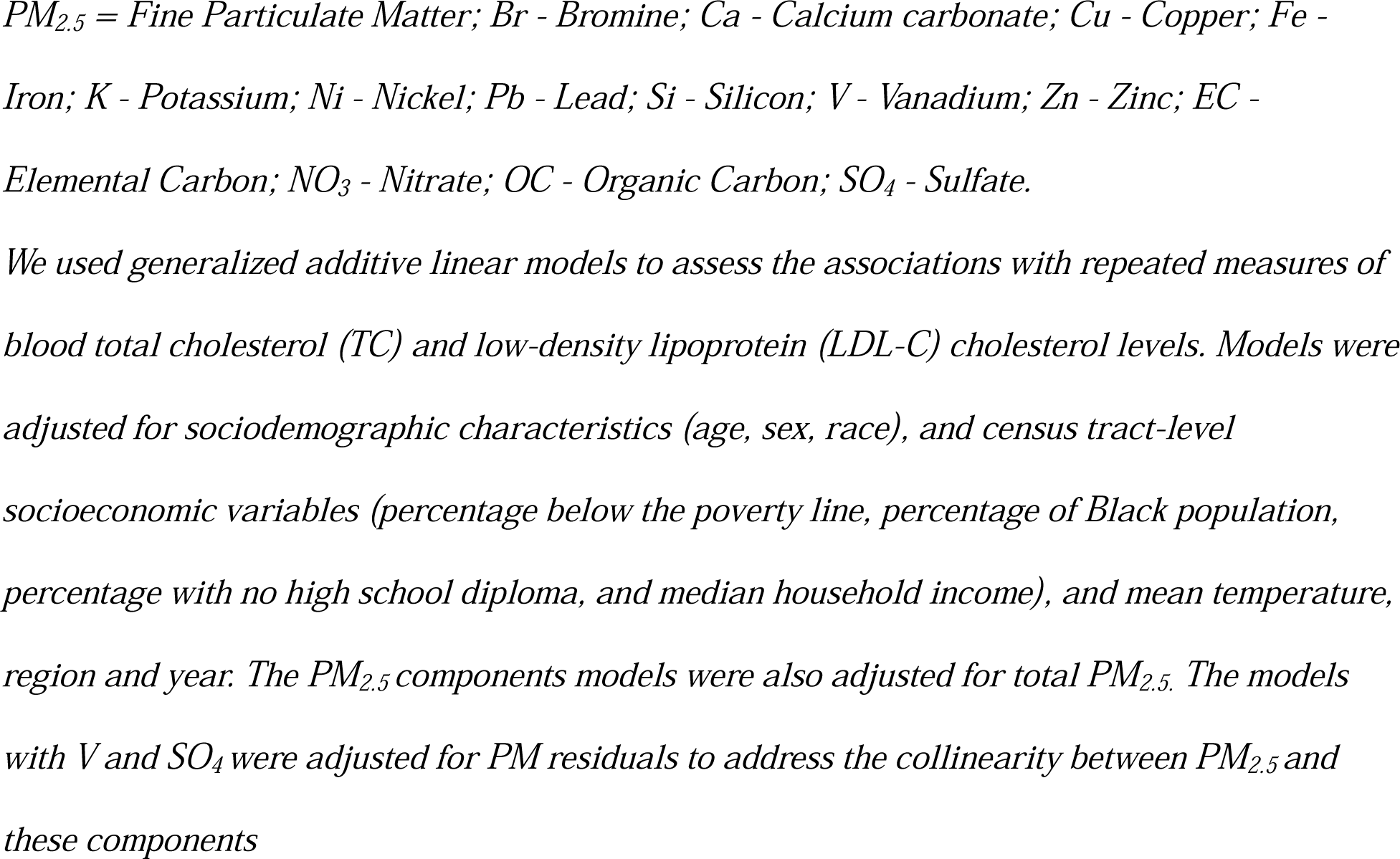
The change in TC and LDL-L associated with interquartile increase in exposures to total and component-specific PM_2.5_: Results of single exposure models.

| Exposure | Change (95% CI) |  |
| --- | --- | --- |
|  | TC | LDL-C |
|  | (N = 143,555) | (N = 96,759) |
| Total PM<sub>2.5</sub><br>(µg/m<sup>3</sup>) | 1.294 (0.579, 2.010) * | 0.827 (0.352, 1.303) * |
| PM<sub>2.5</sub> components |  |  |
| Br | -1.555 (-2.535, -0.574) | -1.205 (-2.512, 0.102) |
| Ca | 0.839 (0.435, 1.242) * | 0.618 (0.187, 1.049) * |
| Cu | 0.921 (0.441, 1.401) * | 0.621 (0.147, 1.095) * |
| Fe | 1.081 (0.630, 1.532) * | 0.748 (0.318, 1.178) * |
| K | -0.205 (-0.578, 0.167) | -0.152 (-0.570, 0.267) |
| Ni | -0.011 (-0.222, 0.201) | 0.363 (0.026, 0.700) * |
| Pb | 0.895 (-0.184, 1.975) | 0.788 (-0.687, 2.263) |
| V | 0.829 (0.134, 1.524) * | 0.874 (0.279, 1.470) * |
| Si | -0.068 (-0.394, 0.258) | -0.110 (-0.387, 0.166) |
| Zn | 0.770 (0.361, 1.179) * | 0.320 (-0.086, 0.725) |
| EC | 0.507 (0.084, 0.930) * | 0.130 (-0.232, 0.492) |
| NO<sub>3</sub> | 0.680 (0.181, 1.180) * | 0.305 (-0.126, 0.736) |
| OC | 0.472 (0.027, 0.918) * | 0.648 (0.136, 1.160) * |
| SO<sub>4</sub> | -1.822 (-3.234, -0.410) | -0.727 (-2.210, 0.755) |

CI = Confidence Interval; TC= Total Cholesterol; LDL-C = low-density lipoprotein.
$PM_{2.5}$ = Fine Particulate Matter; Br - Bromine; Ca - Calcium carbonate; Cu - Copper; Fe - Iron; K - Potassium; Ni - Nickel; Pb - Lead; Si - Silicon; V - Vanadium; Zn - Zinc; EC - Elemental Carbon; $NO_3$ - Nitrate; OC - Organic Carbon; $SO_4$ - Sulfate.
We used generalized additive linear models to assess the associations with repeated measures of blood total cholesterol (TC) and low-density lipoprotein (LDL-C) cholesterol levels. Models were adjusted for sociodemographic characteristics (age, sex, race), and census tract-level socioeconomic variables (percentage below the poverty line, percentage of Black population, percentage with no high school diploma, and median household income), and mean temperature, region and year. The $PM_{2.5}$ components models were also adjusted for total $PM_{2.5}$ . The models with V and $SO_4$ were adjusted for PM residuals to address the collinearity between $PM_{2.5}$ and these components

When evaluating the overall exposure-mixture associations, we found a decile incremental increase in the exposure mixture to be associated with a 0.375 mg/dL increase in TC (95% CI: 0.174, 0.577) driven mainly by OC, Pb, Ca, and Fe, and 0.302 mg/dL increase in LDL-C (95% CI: 0.063, 0.540) similarly driven by Ni, Fe, Pb, V, K, OC, and NO_3_ (Figure 1). The sensitivity analyses showed consistent associations, and the inference remained the same (Table S1). In an analysis examining interactions between exposure-mixture effects and WTC exposures, associations were consistently larger among responders with higher WTC-exposure index rankings, early site arrival (within the first three days), and those who lost someone. However, only the associations with TC were statistically significantly modified by losing someone (interaction p value = 0.006) and the association with LDL-C was statistically significantly modified by WTC-exposure index (interaction p value = 0.043) (Figure 2 and Table S2).

**Figure 1.**
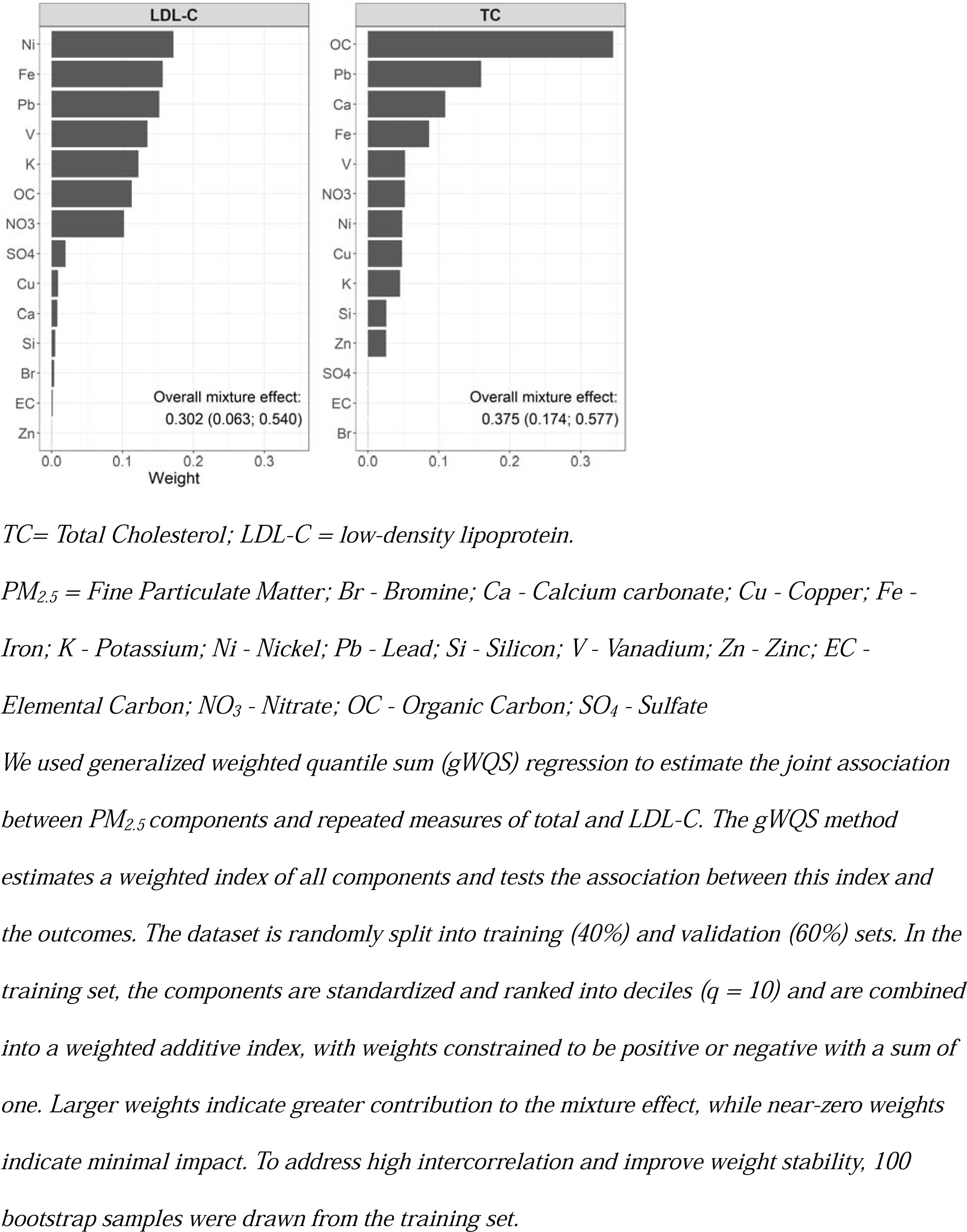
Mixture analysis Weights

**Figure 2.**
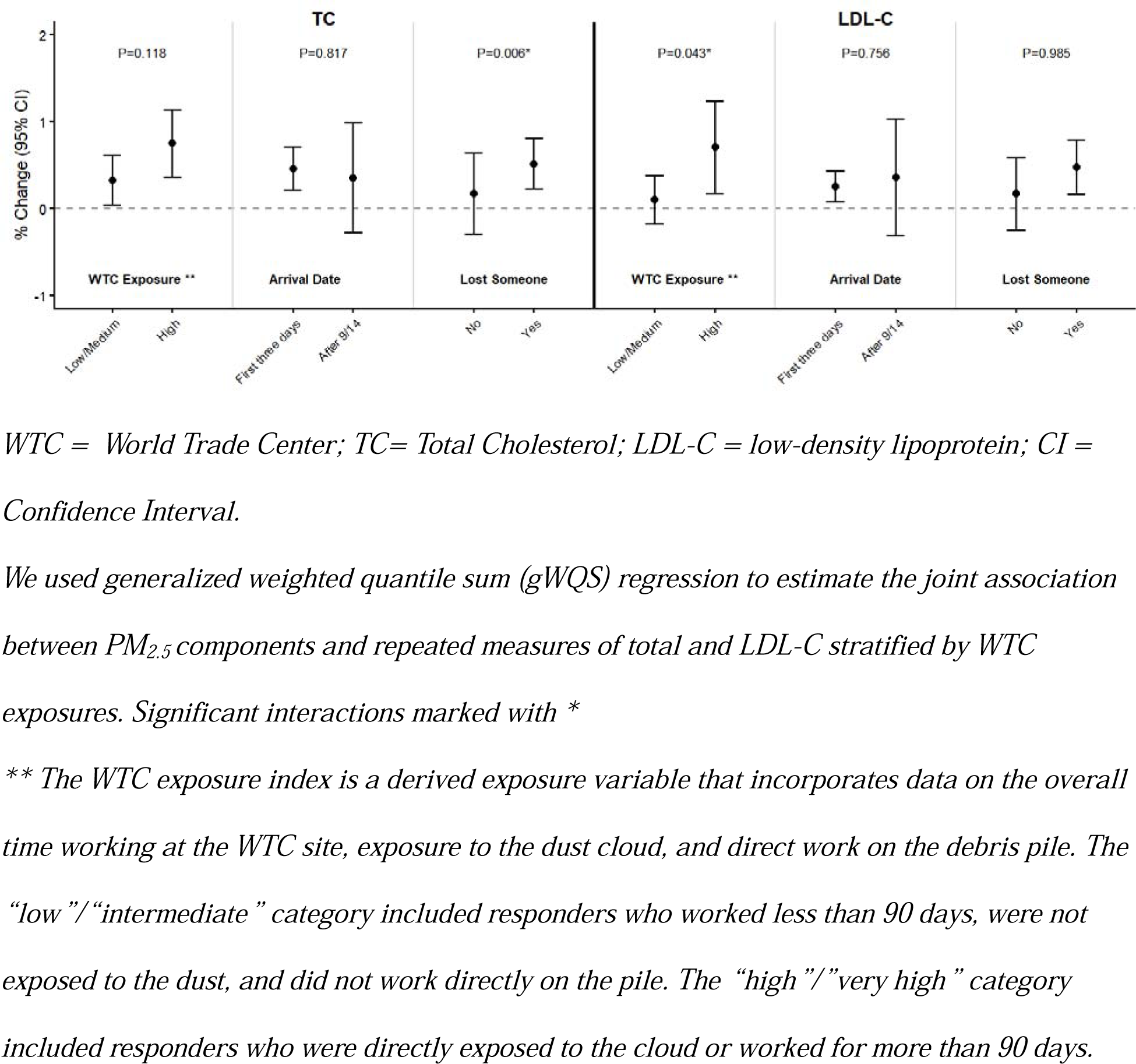
Effect modification of the associations between the exposure mixture with lipids by WTC exposures.

## 4. Discussion

We found consistent associations between PM_2.5_ exposure and elevated lipid levels among WTCHP general responders. This finding is consistent with a previous study that showed that an increase of 0.67 mg/dL (95% CI: 1.00, 2.35) in TC was associated with an IQR increase in PM_2.5_ mass averaged 6 months before the study visit ^2^. However, to the best of our knowledge, currently, there are no other studies linking PM_2.5_ exposure and LDL-C in the WTCHP. While different components (including Pb, Ca, Ni, V, K, and NO_3_) contributed to these associations, organic carbon (OC) and iron (Fe) exposures were consistently found to be major contributors. The associations between PM_2.5_ and TC and LDL-C were previously reported in different meta-analysis studies ^14,15^. One study from China found that 10-μg/m^3^ change in annual PM_2.5_ was linked to a change of 4.2mg/dL in TC and 2.7mg/dL in LDL-C ^16^. A study from the US showed an increase of 2.69% from the mean value (95% CI: 1.87, 3.51) in TC for a 1-μg/m3 increase in annual PM_2.5_ exposure ^17^. Our results suggest that the found 1.294 increase in TC would raise the proportion of lab results above 200 mg/dL from the current 42% to 45%.

Different mechanisms were suggested to explain the relationship between exposure to PM_2.5_ and elevated lipids involving oxidative stress ^18^, mitochondrial dysfunction and reduced absorption ^18^, and stimulated synthesis of related biolipid intermediates inducing elevated LDL-C production ^19^.

PM_2.5_ is a mixture of various chemicals, and its negative effects on blood lipids and dyslipidemia have been reported in previous studies ^14^. We found OC and Fe consistently to be the main drivers of the association between TC and LDL-C. However, studies finding similar associations are very limited. One study in Beijing found an increase in oxidized LDL-C to be associated with short-term (1-day) exposure to iron ^20^. To the best of our knowledge, there are no studies reporting on the associations between OC exposure and lipid levels specifically. Studies reporting on the associations between OC and Fe sources (biomass burning, vehicle emissions, and industrial processes) ^21^ and lipids are focused on biomass burning. For example, a study in Thailand showed that family members of cooks using biomass fuel were at risk of high cholesterol (HC; OR=2.74; 95% CI: 1.66, 4.53) ^22^. This being said, some studies report on the associations with CVD risk. A study in California examined the single-pollutant associations between 19 PM_2.5_ components and daily mortality and found that several constituents (OC, Fe, EC, NO_3_, K, Ti) were associated with CVD deaths at various lags ^23^. Another study found that a higher atherosclerotic CVD mortality rate was associated with oil combustion (1.051 [95 % CI: 1.049, 1.052]), industrial pollution (1.054 [95 % CI: 1.052, 1.056]), coal and biomass burning (1.065 [95 % CI: 1.062, 1.067]), and motor vehicle pollution (1.044 [95 % CI: 1.042, 1.046]) ^24^. Recently, studies using PM_2.5_ mixture analysis are emerging, but with inconsistent results. One study from China found a strong association between the overall effect of PM_2.5_ elemental constituents and TC, with Cu and Ti being the main drivers ^25^. In a different study (also in China), the joint exposure to PM_2.5_ constituents was positively related to dyslipidemia (OR: 1.09, 95 % CI: 1.05–1.14), and the constituent with the largest weight was nitrate (weighted at 0.387) ^6^. A study from Massachusetts, however, found a positive cumulative association between a mixture of components and CVD mortality. They reported that the components with a large contribution to the cumulative associations included K, OC, and Fe ^26^. The variability reported among studies varies by the studied chemical composition, exposure duration, population characteristics, and specific outcome.

Our study also suggests that bromine (Br) and sulfate (SO_4_) exhibit protective associations with TC. Some other studies utilizing chemical composition have found that higher concentrations of Br (sea spray aerosol), and SO_4_ (secondary inorganic aerosol) frequently correlate with a lower risk of elevated health risks compared to more toxic combustion-derived components ^27^. Another study explained that identifying PM_2.5_ constituents such as sulfate as associated with adverse health impacts may be the result of detecting effects of co-varying pollutants (e.g., ammonium sulfate, or other components with similar sources for that region) ^28^.

The differences in the reported significant PM_2.5_ constituents in our study may be a result of the exposure sources, and more importantly, the specific sensitivity of our study population. Several studies have reported on the increased risk of CVD and CVD risk factors in the WTC cohort, highlighting their vulnerability to both acute and later-life environmental exposures ^29,30^. One study demonstrated increases in glucose and cholesterol levels following exposures on 9/11 ^2^, while a subsequent study found that glucose levels also increase in response to later-life exposures ^3^. This hypothesis is supported by our emerging finding that the associations between the exposure mixture and lipid levels were amplified among responders with more intense WTC-related exposures. This was most pronounced for TC, where significantly larger mixture associations were observed among those who lost someone during the 2001 attack and LDL-C among those with a higher WTC-exposure index. The association with TC extends the well-documented link between trauma (specifically Post-traumatic Stress Disorder) and elevated TC levels ^31^. Additionally, its is supported by studies reporting on the high prevalence of PTSD in this cohort ^32,33^. The association between the WTC exposure index and LDL-C should be interpreted cautiously, as it may be influenced by the observed association between WTC exposure and higher body mass index in this cohort ^33,34^. Additionally, previous studies, such as one from 2018 ^33^, did not find these associations significant. However, that study only evaluated WTCHR enrollees who resided in New York City.

The findings from the current study, showing an increase in TC and LDL-C levels following later in life exposures, add further evidence demonstrating increases in cardiovascular-related biomarkers that may translate into elevated CVD risk in this population.

Our findings should be interpreted alongside several limitations. First, although measurement error is a risk in any comprehensive modeling approach, we minimized this potential bias through the high spatiotemporal resolution of our exposure data. Additionally, while we did not adjust for lifestyle factors such as diet and physical activity, our results provide a critical foundation for understanding how environmental stressors independently affect lipid levels. A significant strength of this study is our use of multi-pollutant mixture analysis combined with high-resolution residential exposure data, allowing for a more profound understanding of how combined environmental exposures affect individual lipid profiles.

## 5. Conclusion

In this study we show that higher TC and LDL-C levels are associated with interquartile range increases in both total PM_2.5_ as well as PM_2.5_-specific components such as organic carbon and iron exposure. Higher lipid levels are associated with an increased risk of CVD. Although guidelines provide recommended lipid ranges, lower lipid levels are generally considered a priority. Since CVD is influenced by multiple factors, even a slight increase in lipid levels may contribute to an increased overall risk of developing CVD.

Our findings carry an especially significant weight for the WTCHP responders. We show that responders who experienced more intense WTC-related exposures may be more vulnerable to the effects of later-life environmental exposure mixtures on lipid profiles and CVD risk factors. Therefore, understanding lipid alterations is another step towards clarifying the CVD related consequences in this population.

## Data Availability

The WTC cohort data was obtained from the World Trade Center Health Program (WTCHP) General Responder Cohort (GRC) data center. It includes protected health information with identifiers of individual patients. Investigators wishing to request data can contact the WTCHP GRC data center directly.

## Acknowledgments

Funding and Assistance: This study was supported by the Centers for Disease Control and Prevention, National Institute for Occupational Safety and Health (cooperative agreements and contracts 200-2002-00384, U01-OH008216/23/ 25/32/39/75, 200-2011-39356/61/77/84/85/88,200-2017-93325 and 75D30122C15187). This study was also supported by the Mount Sinai Transdisciplinary Center On Early Environmental Exposures (grant P30 ES023515) and the National Institute of Environmental Health Sciences (grants R01ES034864, KL2TR004421).

## Data statement

The WTC cohort data was obtained from the World Trade Center Health Program (WTCHP) General Responder Cohort (GRC) data center. It includes protected health information with identifiers of individual patients. Per our data use agreement, data supplied for projects with IRB approval shall not be re-used or re-disclosed. Investigators wishing to request data can contact the WTCHP GRC data center directly.

## ABBREVIATIONS

PM_2.5_: Fine Particulate Matter
TC: Total Cholesterol
LDL-C: Low-Density Lipoprotein Cholesterol
CI: Confidence Interval
WTC: World Trade Center
CVD: Cardiovascular Disease
WTCHP: World trade Center Health Program
GRC: General Respondents Cohort
XGBoost: Extreme Gradient Boosting
IDW: Inverse Distance Weight Interpolation
gWQS: generalized weighted quantile sum
BIC: Bayesian Information Criterion

