## Supplementary material for "Long-term Exposure to PM_2.5_ Components and Lipid Profiles in World Trade Center Health Program General Responders": Supplemetary material

**Table S1.** Results from gWQS Mixture models sensitivity analyses

| Exposure | | Change (95% CI) | |
| --- | --- | --- | --- |
|  | TC  (N = 143,555) | | LDL-C  (N = 96,759) |
| Q=10 | 0.375 (0.174; 0.577) | | 0.302 (0.063; 0.540) |
| Q=9 | 0.421 (0.129; 0.713) | | 0.306 (0.038; 0.574) |
| Q=8 | 0.465 (0.217; 0.713) | | 0.378 (0.034; 0.722) |

*CI = Confidence Interval; TC= Total Cholesterol; LDL-C = low-density lipoprotein*

*As a sensitivity analysis, we repeated the generalized weighted quantile sum (gWQS) regression using different quartiles instead of deciles to assess the robustness of the mixture effect and component weights to the choice of quantile specification*

**Table S2.** Effect modification of the associations between the exposure mixture with lipids by WTC exposures.

|  | % Change (95% CI) | |
| --- | --- | --- |
| Modifier | TC | LDL-C |
| WTC exposure index ** |  |  |
| *Low/Medium* | 0.323 (0.035; 0.612) * | 0.099 (-0.177; 0.376) |
| *High* | 0.750 (0.362; 1.138) * | 0.705 (0.176; 1.234) * |
| *Interaction p value* | 0.118 | 0.043 * |
| Arrival date |  |  |
| *First three days* | 0.457 (0.209; 0.705) * | 0.254 (0.073; 0.435) * |
| *After 9/14* | 0.354 (-0.279; 0.988) | 0.358 (-0.309; 1.027) |
| *Interaction p value* | 0.817 | 0.756 |
| Lost Someone |  |  |
| *No* | 0.171 (-0.293; 0.635) | 0.169 (-0.247; 0.587) |
| *Yes* | 0.513 (0.222; 0.803) * | 0.476 (0.164; 0.787) * |
| *Interaction p value* | 0.006 * | 0.985 |

*WTC = World Trade Center; CI = Confidence Interval; TC= Total Cholesterol; LDL-C = low-density lipoprotein.*

*** The WTC exposure index is a derived exposure variable that incorporates data on the overall time working at the WTC site, exposure to the dust cloud, and direct work on the debris pile. The “low”/“intermediate” category included responders who worked less than 90 days, were not exposed to the dust, and did not work directly on the pile. The “high”/”very high” category included responders who were directly exposed to the cloud or worked for more than 90 days.*

**Figure S1.** The associations between PM_2.5_ components and TC (a) and LDL-C (b) using penalized splines of the exposures.

a

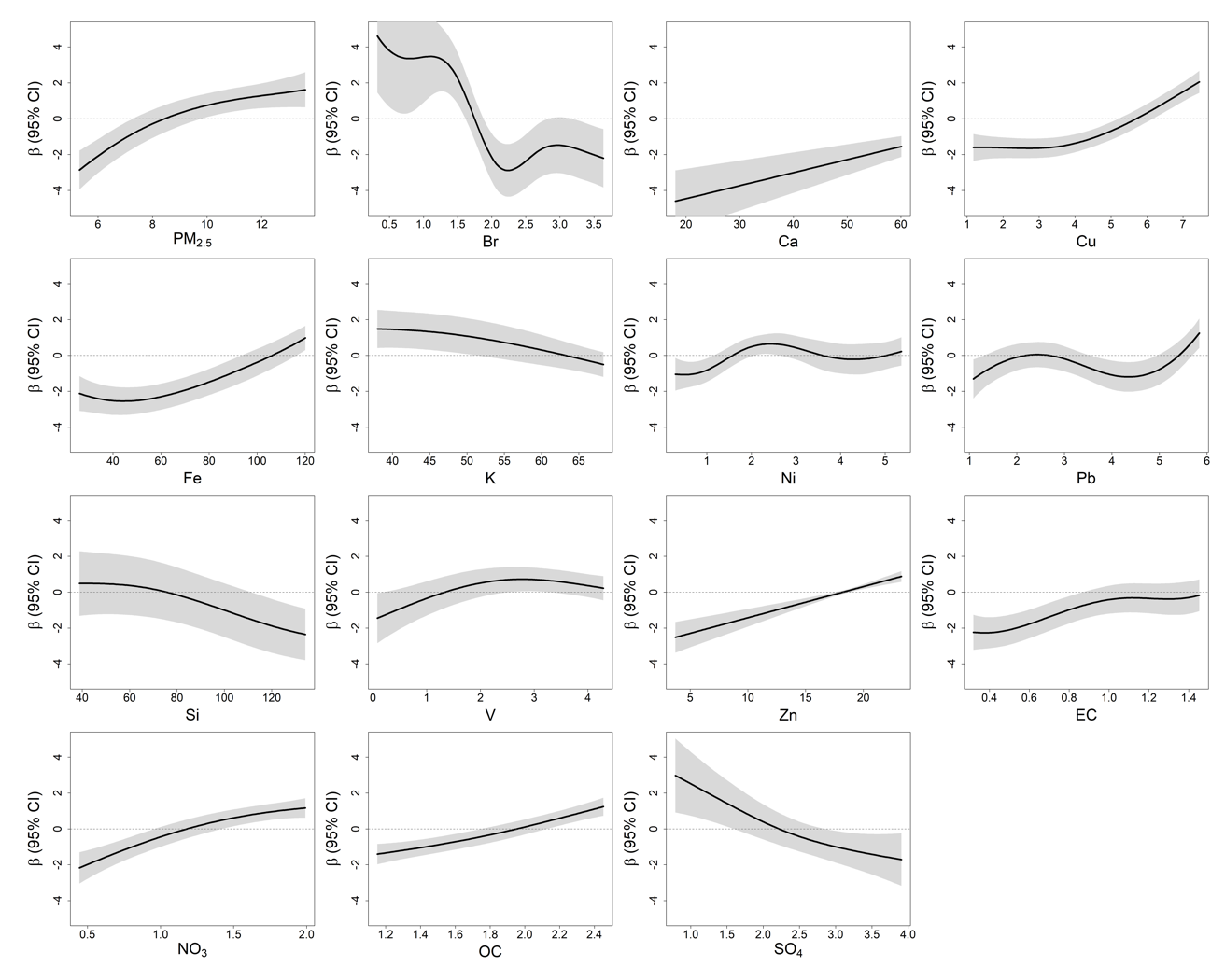

b

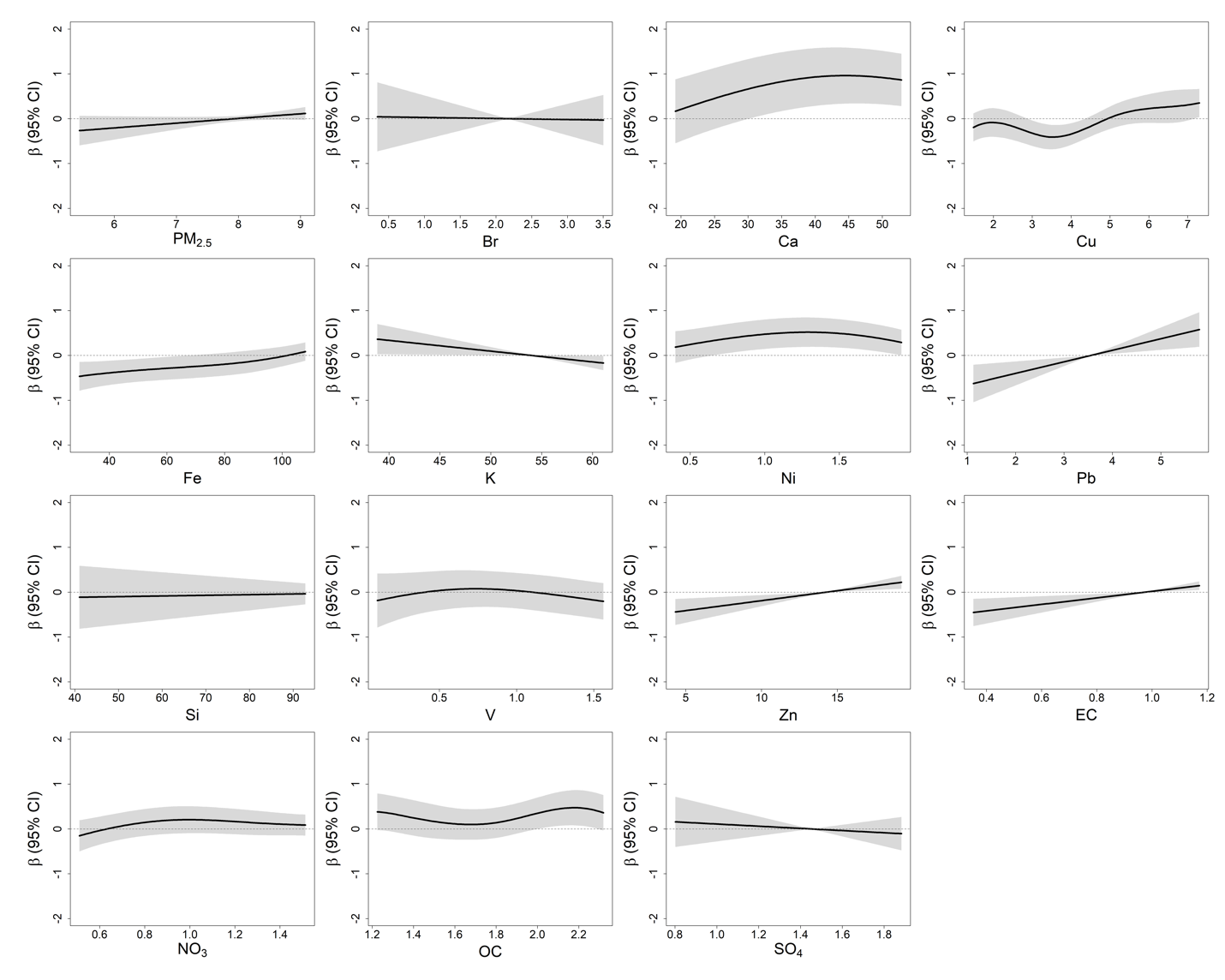

*PM_2.5_ = Fine Particulate Matter; CI = Confidence Intervals;*

*TC= Total Cholesterol; LDL-C = low-density lipoprotein.*

*Br - Bromine; Ca - Calcium; Cu - Copper; Fe – Iron; K - Potassium; Ni - Nickel; Pb - Lead; Si - Silicon; V - Vanadium; Zn - Zinc; EC - Elemental Carbon; NO_3_ - Nitrate; OC - Organic Carbon; SO_4_ – Sulfate.*

*All smooth terms were plotted using a common fixed y-axis scale to facilitate visual comparison across PM_2.5_ components. Exposure-response curves were restricted to the 5th-95th percentile range of each component to avoid instability and sparse data at the distribution tails. Shaded areas represent 95% confidence intervals, with the horizontal dashed line indicating the null (β = 0).*

**Figure S2.** The correlations between the studied PM_2.5_ components.

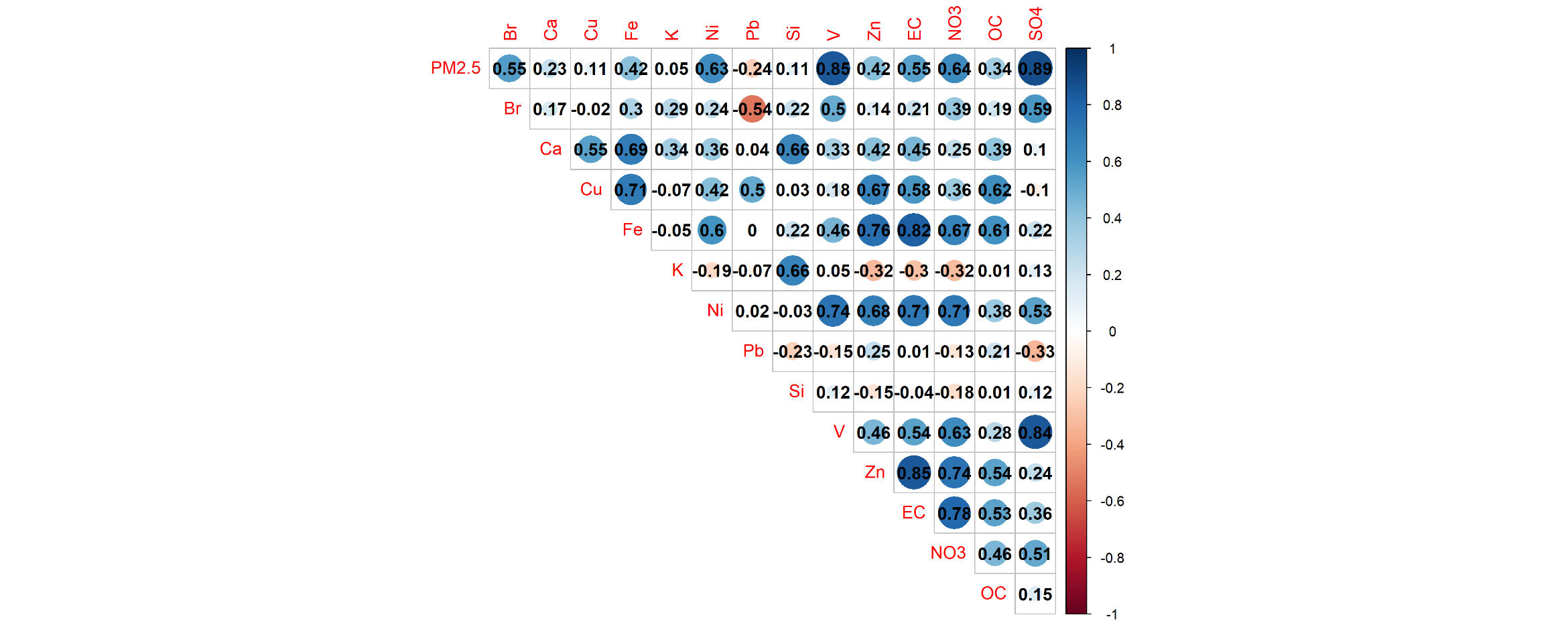

*PM_2.5_ = fine particulate matter; Br - Bromine; Ca - Calcium Carbonate; Cu - Copper; Fe - Iron; K - Potassium; Ni - Nickel; Pb - Lead; Si - Silicon; V - Vanadium; Zn - Zinc; EC - Elemental Carbon; NO_3_ - Nitrate; OC - Organic Carbon; SO_4_ - Sulfate.*
